# Comparative Analysis of Nebulizers in Clinical use for Pressurized Intraperitoneal Aerosol Chemotherapy (PIPAC)

**DOI:** 10.1101/2023.03.24.23287646

**Authors:** Daniel Göhler, Kathrin Oelschlägel, Mehdi Ouaissi, Urs Giger-Pabst

## Abstract

**Objective:** Technical ex-vivo comparison of commercial nebulizer nozzles used for Pressurized Intraperitoneal Aerosol Chemotherapy (PIPAC).

**Methods:** The performance of four different commercial nebulizer nozzles (Nebulizer; HurriChem™; MCR-4 TOPOL^®^; QuattroJet) was analysed by comparing: i) technical design and principle of operation, ii) operational pressure as function of the liquid flow rate, iii) droplet size distribution via laser diffraction spectrometry, iv) spray cone angle, spray cone form, and horizontal drug deposition through image-metric analyses, and v) chemical resistance via exposing to a cytostatic solution and metallurgic composition by means of spark optical emission spectral analysis.

**Results:** The Nebulizer exhibits a nearly identical technical design, implying a comparable performance (e.g., mass median droplet size of 29 μm) as the original PIPAC nozzles (MIP/ CapnoPen). The other three nozzles demonstrate varying degrees of performance deviation from the original PIPAC nozzles. The HurriChem™ shares a similar design and principle of operation as the Nebulizer, but produces a finer aerosol with a particle size of 22 μm. The operating principles of MCR-4 TOPOL^®^ and QuattroJet significantly differ from that of the original PIPAC nozzle technology. The MCR-4 TOPOL^®^ nebulizer has a hollow spray cone that leads to the production of significantly larger aerosol droplets (50 μm) compared to the original PIPAC nozzles. The QuattroJet generates an aerosol droplet (22 μm) similar in size to the HurriChem™ and exhibits improved spatial drug distribution.

**Conclusion:** While the introduction of new PIPAC nozzles is a welcome development, differences in performance and efficacy were noted. Therefore, it is recommended that PIPAC nozzles that deviate from the current standard undergo bioequivalence testing and be implemented in accordance with the IDEAL-D framework prior to routine clinical use.

## 1 Introduction

More than a decade ago, Pressurized Intraperitoneal Aerosol Chemotherapy (PIPAC) was introduced clinically as a new approach to deliver intraperitoneal chemotherapy to patients suffering from end-stage peritoneal surface malignancies. Using a high-pressure injector connected to a specially designed PIPAC nozzle, liquid chemotherapeutic drugs are aerosolised during laparoscopic surgery within the capnoperitoneum. This approach is expected to have a better spatial drug distribution pattern, greater depth of tissue penetration, and higher drug concentration in the tissue than conventional liquid intraperitoneal chemotherapy [1, 2]. Clinical data from phase I/II and larger mono- and multicentre case series regarding safety, feasibility, and oncologic efficacy are encouraging. While the therapeutic role of PIPAC remains uncertain [3], there are ongoing prospective randomized PIPAC trials, and their results are eagerly awaited [4, 5].

Until recently, the original PIPAC nozzle was the only option available for clinical use. More than 18’000 documented clinical applications worldwide were recorded by the end of 2022 [3]. Significant efforts have been dedicated to standardizing PIPAC therapy globally in order to compare efficacy. [6, 7]. Technical and clinical performance of the original PIPAC nozzle have been extensively studied in pre- and clinical settings [3, 8, 9]. The recent introduction of new nebulizer devices and their clinical use requires a detailed evaluation of their physical properties. As of now, very limited comparative data are available for the newly introduced PIPAC nozzles. Oncological surgeons are now confronted with the question of whether these newer nozzles are equivalent to the original nozzle technology or if they possess potential technical or functional advantages or disadvantages that could impact oncological outcomes.

Building upon the methodological findings regarding the technical characterization of the original PIPAC nozzle [8], the present study focuses on the comparative performance characterization of four commercially available nebulizers commonly used in PIPAC procedures.

## 2 Materials and Methods

### 2.1 Examined PIPAC nozzles

Four commercial single-substance PIPAC nozzles for intraperitoneal drug aerosolization were examined, i.e.,

- Nebulizer, Model 770-12, REGER Medizintechnik, Villingendorf, Germany (A),
- HurriChem™, ThermaSolutions, White Bear Lake, MN, United States of America (B),
- MCR-4 TOPOL^®^, SKALA-Medica, Soběslav, Czech Republic (C),
- QuattroJet, Model 770-14, REGER Medizintechnik, Villingendorf, Germany (D).

After the experiments, all nozzles were longitudinally cut open in the middle at a 180° angle using a computerized numerical control milling machine to investigate their principles of operation. In addition, also the dimensions of the nozzle outlet orifices were examined by light microscopy (SMZ1500, Nikon, Tokyo, Japan).

### 2.2 Barometric characterisation of operational pressure as function of liquid flow rate

To characterise the operational pressure over the volumetric liquid flow rate, the nozzles were connected via high-pressure hose lines with a high-pressure injector (ACCUTRON^®^ HP-D, MEDTRON AG, Saarbrücken, Germany) to push the test liquid (Glucosterile 5%, Fresenius Kabi GmbH, Germany) through the nozzles. The operational pressure induced by the liquid flow rate was determined by means of a glycerine-filled bourdon gauge (MA7U-25, JRA Maschinenteile und Geräte GmbH, Reichenbach, Germany), which was implemented in the high-pressure line. For the analyses, the volumetric liquid flow rate was increased stepwise either by 0.1 ml/s (for nozzles A, B and D) or by 0.2 ml/s (for nozzle C) until the maximum permitted pressure of 21 bar of the high-pressure injector was reached. For nozzle D, only the axial nozzle was tested - the horizontal nozzles were sealed watertight. Analogous to [8], measurement values were taken at steady state conditions of the aerosolization process, and all analyses were repeated three times.

### 2.3 Granulometric characterisation of droplet size distributions

The droplet size distributions of the aerosols generated from the test liquid (Glucosterile 5%, Fresenius Kabi GmbH, Germany) were characterised by laser diffraction spectrometry (PW180-C spray particle size analyser, Jinan K-Ring Technology Co., Ltd, Shandong, China) over a size range of (0.57 - 780) μm. The outlets of the PIPAC nozzles were arranged via a tripod in a distance of 5 mm perpendicular to the centre of the free-accessible red laser beam. To characterise the aerosolization performance, all analyses were performed contemporaneous with the barometric characterisation of the operational pressure for various liquid flow rates. Analogous to [8], measurement values were taken at steady state conditions of the aerosolization process and all analyses were repeated three times.

### 2.4 Image-metric characterisation of spray cone angles, form and horizontal drug deposition areas

The spray cone angles, the form of the spray cones and the horizontal drug deposition area were characterised with different test liquids at nozzle-specific operation conditions as recommended by the manufacturers, i.e., at a volumetric liquid flow rate of 0.5 ml/s for the nozzles A, B, at 2.0 ml/s for nozzle C and 1.5 ml/s for nozzle D. The former two characteristics were evaluated on the base of a 5 wt.-% aqueous glucose solution (Glucosterile 5%, Fresenius Kabi GmbH, Germany), while the latter characteristic was assessed by operating the nozzles with undiluted royal blue ink (Pelikan Tinte 4001^®^, Hannover, Germany).

For the spray cone angle analyses, the nozzles were fixed on a tripod and vertically aligned. Photographic images were taken with a camera that was perpendicular positioned to the nozzle direction. The images were in-silico processed by overlaying with a digital 360° full-circle protractor for determining the spray cone angles.

The form of the spray cones was visualized by means of a line laser (GCL 2-15, Robert Bosch Power Tools GmbH, Leinfelden-Echterdingen, Germany) positioned in distance of 60 mm from the nozzle orifice at right angle into the spray cone. Fully evaluated spray cone forms were finally documented photographically.

The horizontal drug deposition on a level-aligned blotting paper was examined by operating the vertically aligned nozzles with a distance of 60 mm between the blotting paper and the nozzle orifice. The blotting paper was exposed for 3 s to the fully-developed spray jet. To achieve this, a mechanical diaphragm was placed in front of the spray jet. The diaphragm was opened automatically within 0.1 s, when the aerosol jet showed steady state nebulisation condition.

### 2.5 Assessing of chemical resistance and chemical composition

To assess the chemical resistance of the nozzle material against chemotherapeutic drugs, the nozzles were at first exposed to a cytostatic solution for 12 hours and afterwards stored in the dark at room temperature for 12 days within petri dishes. The chosen cytostatic solution was prepared in accordance to the mixture of high pressure/high dose PIPAC (HP/HD-PIPAC) [10], i.e., 6 mg of doxorubicin (Accord 2 mg/ml, Accord Healthcare GmbH, Munich, Germany) was admixed to a total volume of 50 ml with a 0.9 wt.-% aqueous sodium chloride solution (Ecolav^®^ 100, B. Braun, Melsungen, Germany). Finally, the nozzles were milled open in a laminar flow workbench and macroscopic changes were documented photographically.

Furthermore, the chemical composition of the nozzles pipes was characterised for the following elements: C, Si, Mn, P, S, Cr, Ni, Mo, Cu, W and N by means of spark optical emission spectral analysis (SPECTROMAXx, SPECTRO Analytical Instruments GmbH, Kleve, Germany) via an independent, state-recognized laboratory (WS Material Service GmbH, Essen, Germany).

## 3 Results

### 3.1 Technical design and principle of operation

The 90° sectional views of the head regions in Figure 1 show technical destails of the examined nozzles.

**Figure 1:**
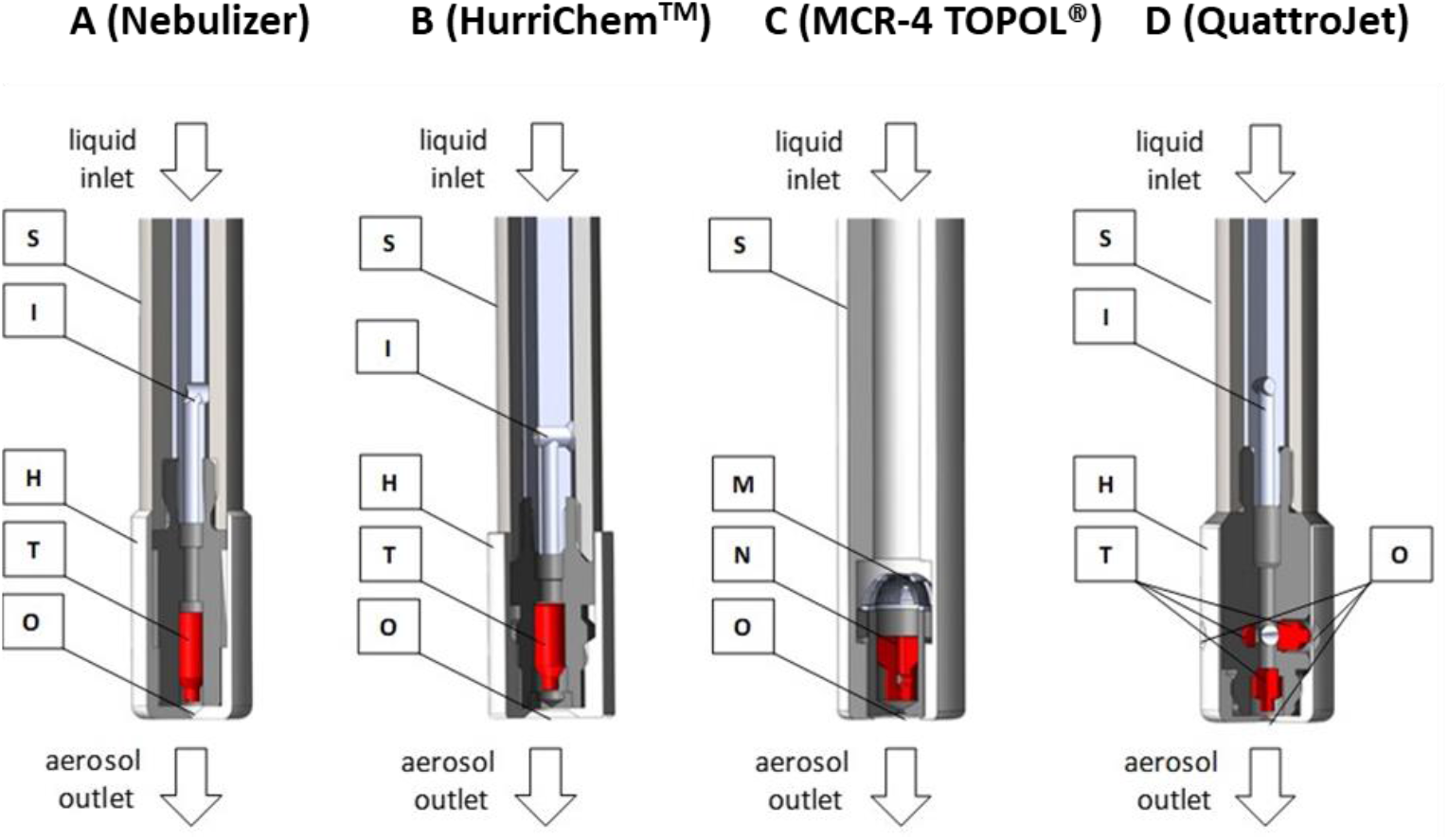
90° sectional views of the head regions of the nozzles. Legend: O = outlet orifice; H = nozzle head; I = bar inlay with distal transverse borehole; M = double metal grid; N = fixed needle; S = shaft; T = twist body.

Externally, all nozzles consist of a stainless steel shaft (S) with a more or less pronounced nozzle head (H) on the lower part and a Luer lock thread on the upper part (not shown in Figure 1). The Luer lock threads serve for the connection of the nozzles with high-pressure injectors via high-pressure hose lines. Internally, the nozzles show considerable differences (Figure 1). It is worth noting that nozzle A and B exhibit a nearly identical construction and principle of operation. However, both nozzle C and D differ significantly from each other and from nozzle A and B

In the case of the nozzles A, B and D, the liquid drug is supplied internally from the Luer lock connector to the nozzle head via an annular gap between the outer shaft (S) and a bar inlay with distal transverse borehole (I). In contrast, the internal liquid drug supply of nozzle C occurs directly via the hollow cavity of the shaft (S). Moreover, nozzle C is equipped with a double metal grid (M) with two different mesh sizes that serve as particle filter.

While the nozzles A and B contain one twist body (T), nozzle D is equipped with four twist bodies (i.e., with one axial and three lateral twist bodies in 120° arrangement) to improve the spatial drug distribution within the abdominal cavity. The twist bodies (T) of the nozzles A, B and D contain longitudinally superfically milled grooves at 180° intervals. As the liquid drug flow rate passes along the twist bodies (T) they were set into rotation that improves the aerosolisation prior leaving the nozzle via the outlet orifice (O). In the case of nozzle C, the twist body is replaced by an fixed metal needle (N). This needle contains also laterally located, spirally milled axial grooves that induce a whirlwind effect for aerosolisation when passed by the liquid flow before leaving the nozzle via the outlet orifice (O).

Light microscopic images of the oulet orifices (O) with determined orifice diameters of the examined nozzles are shown in Figure 2.

**Figure 2:**
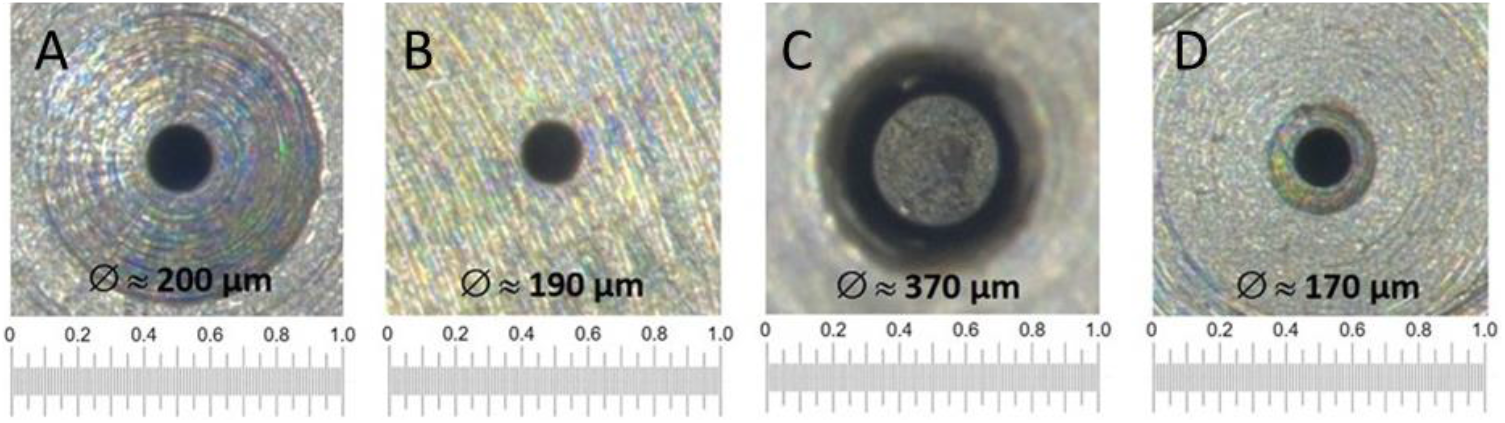
Light microscopic images of the outlet orifices with determined orifice diameters of the examined nozzles; scaling in mm. Nozzles A, B, C, D

### 3.2 Operational parameters based on barometric and granulometric analyses

In Figure 3a, the operational pressure of the examined nozzles is shown as a function of the liquid flow rate. Figure 3b displays the mass median diameter of the droplet size distribution over the operational pressure. To avoid artefacts due clouding of the optics of the laser diffraction spectrometer, the lateral nozzles of nozzle D (QuattroJet) were taped off for the granulometric analyses and a flow rate of 0.5 ml/s was chosen (manufacturer-recommended flow rate of 1.5 ml/s) Note that the shown data are determined at steady state conditions of the aerosolization process analogous to [8]. Figure 3a shows that the determined operational pressure data for all examined nozzles fit well with the fluid dynamic theory, i.e., the dynamic pressure (or the dynamic pressure drop) of an incompressible fluid increases with the fluid velocity by the power of two. According to the equation of continuity, the fluid velocity of an incompressible fluid is in turn directly proportional to the volumetric liquid flow rate. The nozzles A, B and D show a similar performance regarding operational pressure and liquid flow rate, while nozzle C (MCR-4 TOPOL^®^) has a significantly lower pressure drop, and thus, a considerable higher volumetric liquid flow rate at a specific operational pressure.

**Figure 3:**
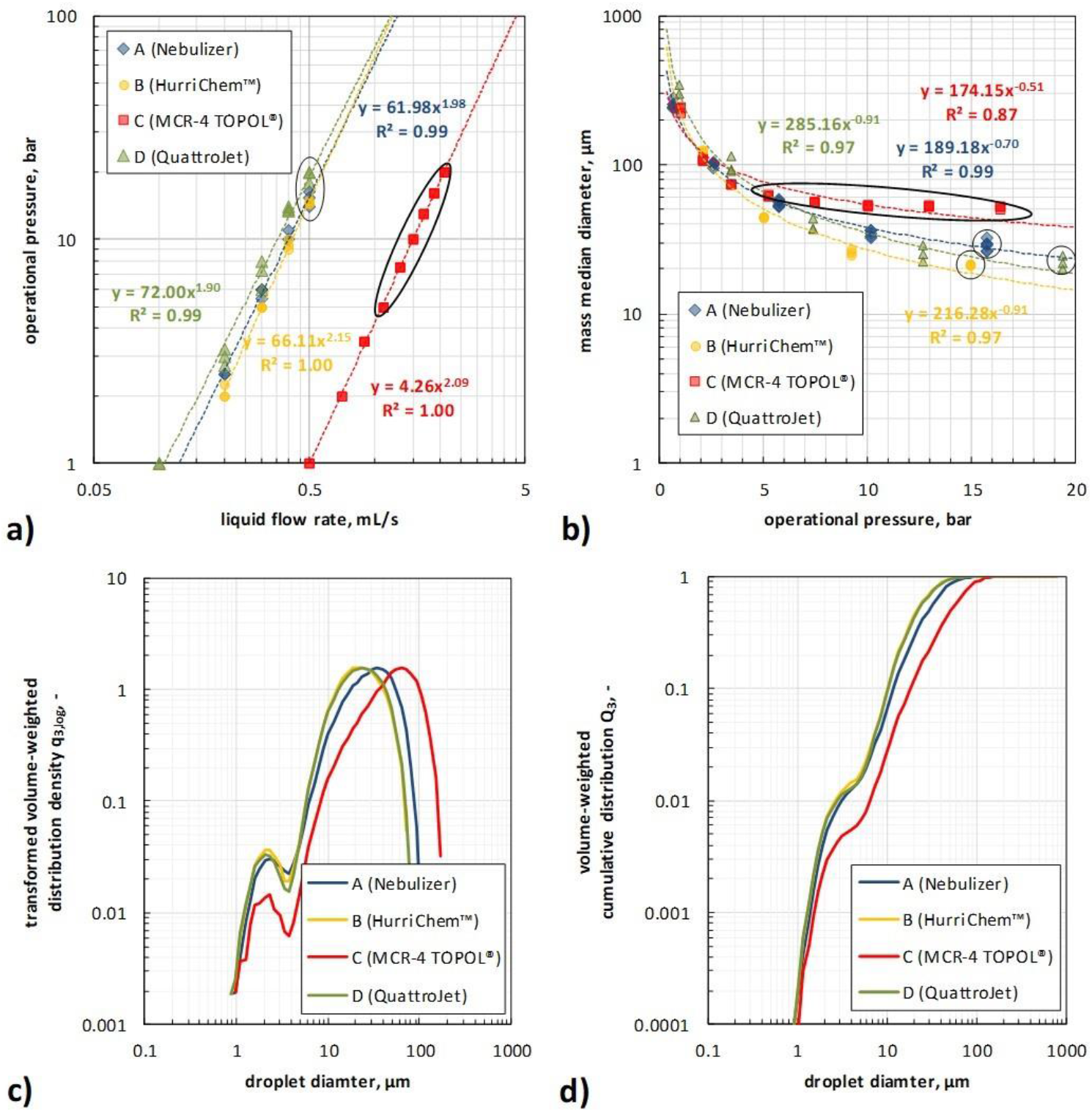
Operational pressure as function of the liquid flow rate from barometric analyses (a), mass median diameter as function of the operational pressure from granulometric analyses (b) and volume-weighted distributions density (c) and cumulative distribution (d) of droplets at certain manufacturer-recommended operational condition; black cycles/ellipses indicate manufacturer-recommended operation condition.

In addition to the entire operational spectrum, the study also separately investigated the manufacturer-recommended operational conditions, i.e., at a volumetric liquid flow rate of 0.5 ml/s for nozzle A (Nebulizer) and nozzle B (HurriChem™), 1.3 - 2.0 ml/s for nozzle C (MCR-4 TOPOL^®^) and 1.5 ml/s for nozzle D (QuattroJet). Under these conditions nozzle C showed with (18 - 26) s the shortest initiation time to reach the corresponding steady state pressure of (7.4 - 18.1) bar, followed by nozzle A with 52 s (15.7 bar) and nozzle D with 94 s (16.0 bar). Note that with taped-off lateral nozzles, nozzle D shows a higher operational pressure of 19.3 bar (as shown in Figure 3).

Figure 3b shows that the mass median diameter of the generated droplet aerosols depends for each nozzle significantly on the operational pressure. With increasing operational pressure, the mass median diameter decreases. For operational pressures of ≤ 4 bar, no significant differences between the different nozzles were observed. This is attributed to a non-fully developed aerosolization of the supplied liquid. For operation pressures of ≥ 5 bar stable aerosol generation is reached and differences between the nozzles can be observed. For operational pressures ≥ 5 bar, nozzle C (MCR-4 TOPOL^®^) shows the coarsest mass median diameters, followed by nozzle A (Nebulizer). The finest mass median diameters were determined for nozzle B (HurriChem™) and D (QuattroJet).

The same ranking can also be deduced by the volume-weighted droplet size distributions of the aerosols as generated by the nozzles at the manufacturer-recommended operation conditions (Figure 3c, Figure 3d). Moreover, it can be observed in Figure 3c and Figure 3d, that each aerosol has a polydisperse and bimodal droplet size distribution.

### 3.3 Operational parameters based on image-metric analyses

Figure 4 shows photographic images for the spray cone angle (upper panel), the spray cone form (mid panel) and the horizontal drug deposition area (lower panel) of each examined nozzle as determined at manufacturer-recommended operational conditions.

**Figure 4:**
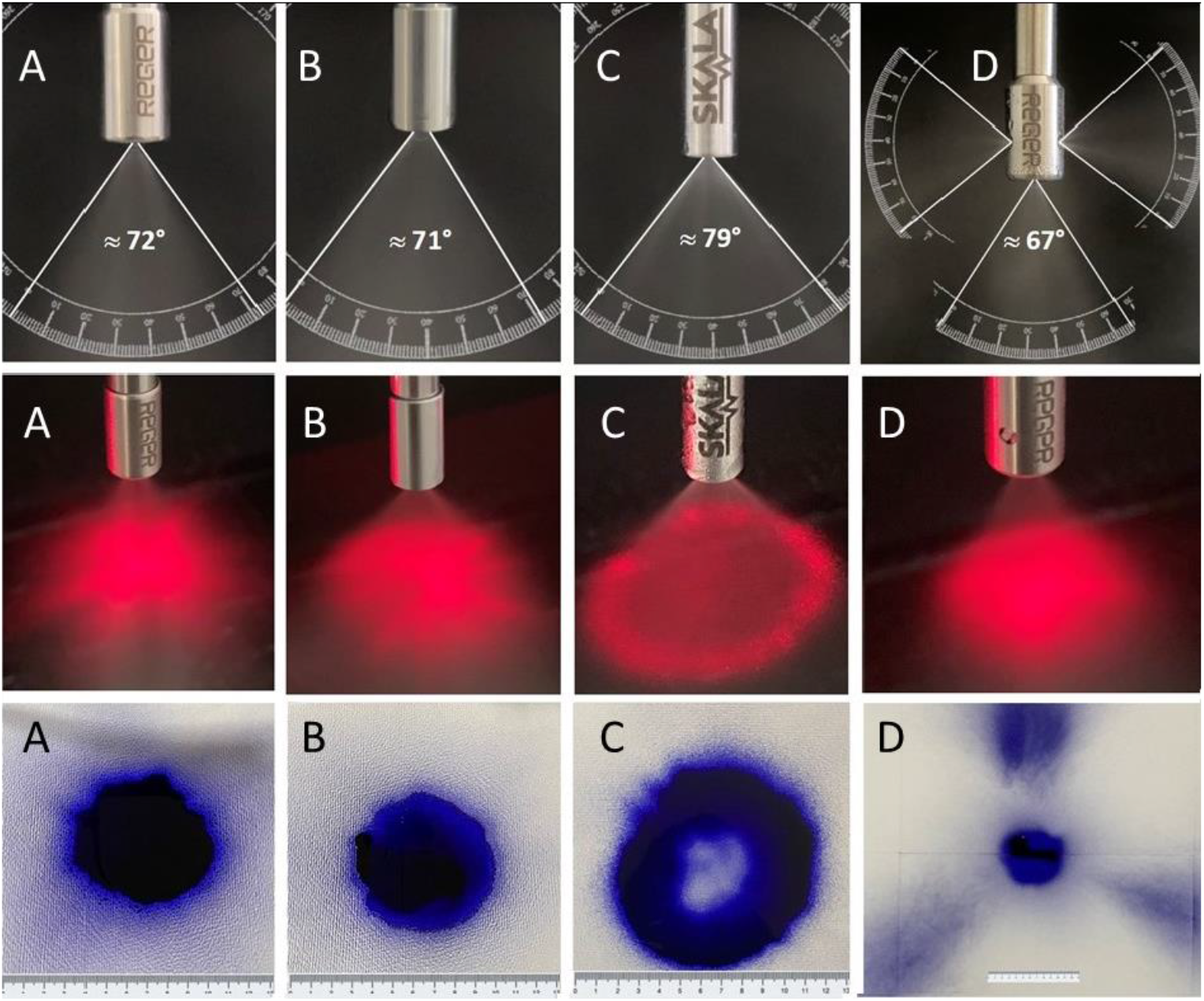
Photographic images of spray cone angle (upper panel), of spray cone form (middle panel) and horizontal drug deposition area (lower panel, scale in cm). Nozzles A, B, C, D.

According to the upper panels of Figure 4, the widest single spray cone angle, 79°, was achieved with nozzle C (MCR-4 TOPOL^®^), 72° for nozzle A (Nebulizer), 71° for nozzle B (HurriChem™), and 67° for nozzle D (QuattroJet). While Nozzle D exhibits the smallest single spray cone angle (67°), it should be noted that unlike the other nozzles, Nozzle D consists of four spray cones. As shown in the middle panels of Figure 4, nozzle A (Nebulizer), nozzle B (HurriChem™) and nozzle D (QuattroJet) generate a full spray cone, whereas nozzle C (MCR-4 TOPOL^®^) produces a hollow spray cone. The full spray cones of the nozzles A, B and C lead to completely filled circular areas of horizontal drug deposition beneath the nozzles as shown in the lower panel of Figure 4. In the case of the nozzles A and B, a circular deposition area of approx. 38.5 cm^2^ (outer diameter of approx. 7 cm) was determined. The lateral outlets of nozzle D showed in addition to the axial circle (outer diameter of approx. 7 cm) 3 additional deposition areas of (13 × 20) cm that accumulate to an overall horizontal deposition area of approx. 679 cm^2^.

### 3.5 Chemical resistance and chemical composition

Photographic images of the nozzle parts after prolonged exposure to the cytostatic solution are shown in Figure 5.

**Figure 5:**
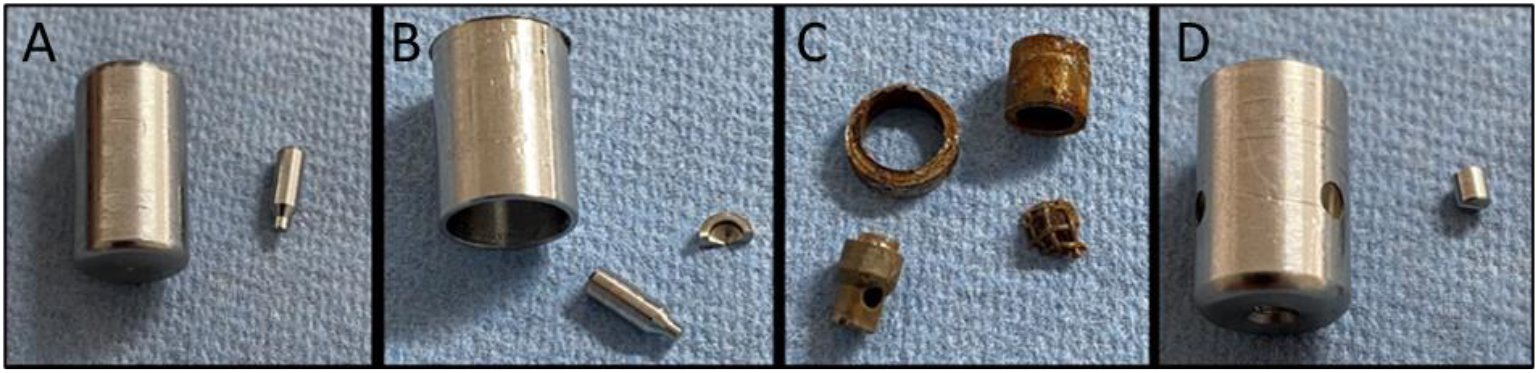
Photographic images of the nozzle parts after exposure to the cytostatic solution. Nozzle A, B, C, D.

As shown in Figure 5, no macroscopic corrosion was observed except for nozzle C (MCR-4 TOPOL^®^). Prolonged exposure to cytostatic solution led to the formation of iron oxide. Corrosion was particularly pronounced on the fine-mesh particle filter, the nozzle needle, and the nozzle head housing. No changes were observed for the nozzles A, B and D either visually or by light microscopic analyses.

Analysis of chemical composition revealed that nozzles A, B and D fulfil requirements of stainless steel 1.4301, typically used for surgical instruments according to EN 10088-3:2014 [11]. Nozzle C (MCR-4 TOPOL^®^) showed a twelve times higher quantity of sulphur (0.012 wt.-% vs. 0.001wt.-%). In addition, molybdenum (0.183 wt.-%), copper (0.220 wt.-%) and tungsten (0.134wt.-%) were identified by spark optical emission spectrometry for nozzle C.

## 4 Discussion

Given the current lack of knowledge, this study conducted a comparative performance analysis of four clinically used nebulizing nozzles for PIPAC. The key technical characteristics of these nozzles are summarised in Table 1.

**Table 1:**
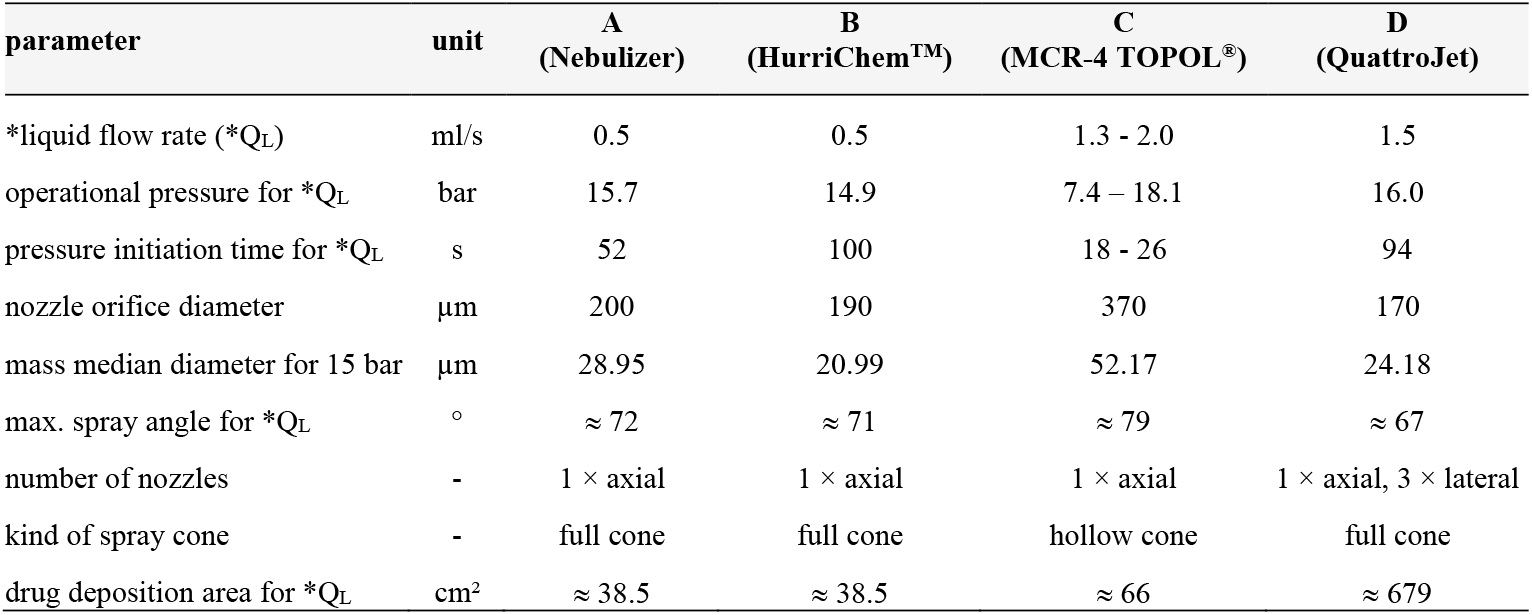
Overview on technical and functional characteristics of the examined nozzles; * = manufacturer-recommended operational conditions.

| parameter | unit | A<br>(Nebulizer) | B<br>(HurriChem™) | C<br>(MCR-4 TOPOL®) | D<br>(QuattroJet) |
| --- | --- | --- | --- | --- | --- |
| *liquid flow rate (*Q<sub>L</sub>) | ml/s | 0.5 | 0.5 | 1.3 - 2.0 | 1.5 |
| operational pressure for *Q<sub>L</sub> | bar | 15.7 | 14.9 | 7.4 – 18.1 | 16.0 |
| pressure initiation time for *Q<sub>L</sub> | s | 52 | 100 | 18 - 26 | 94 |
| nozzle orifice diameter | µm | 200 | 190 | 370 | 170 |
| mass median diameter for 15 bar | µm | 28.95 | 20.99 | 52.17 | 24.18 |
| max. spray angle for *Q<sub>L</sub> | ° | ≈ 72 | ≈ 71 | ≈ 79 | ≈ 67 |
| number of nozzles | - | 1 × axial | 1 × axial | 1 × axial | 1 × axial, 3 × lateral |
| kind of spray cone | - | full cone | full cone | hollow cone | full cone |
| drug deposition area for *Q<sub>L</sub> | cm<sup>2</sup> | ≈ 38.5 | ≈ 38.5 | ≈ 66 | ≈ 679 |

Nozzle A (Nebulizer) shows after an initiation time of 52 s an operational pressure of 15.7 bar at the manufacturer-recommended operational liquid flow rate of 0.5 ml/s. Thereby, a full spray jet cone (71°) composed of droplets with a mass median diameter of 29 μm is formed. The data of this study reveal that nozzle A is identical in design and performance to the nozzle type that was already introduced 10 years ago for clinical use (microinjection pump (MIP)) [8], which was also distributed under the tradename CapnoPen.

Nozzle B (HurriChem™) is another nebulizer that has recently been introduced for PIPAC use. Examinations on design and principle of operation show a high similarity with nozzle A (Nebulizer) and thus also with the initial PIPAC nozzle technology. At the manufacturer-recommended operational liquid flow rate of 0.5 ml/s, nozzle B shows after an initiation time of 100 s an operational pressure of 14.9 bar. The mass median diameter of the droplets in the formed full spray jet cone (73°) was determined to be 21 μm.

Nozzle C (MCR-4 TOPOL^®^) differs from other nozzles investigated in terms of technical design, principle of operation, operational parameters, and aerosol characteristics. The operation of nozzle C is accompanied by the formation of a hollow spray cone jet (79°). At the manufacturer-recommended operational liquid flow rate range of (1.3 - 2.0) ml/s, operational pressures of (7.4 - 18.1) bar were reached rapidly (18 - 26) s. The mass median droplet size decreases with increasing liquid flow rate but was found to be in each case larger than 50 μm.

Nozzle D (QuattroJet) is a further PIPAC nebulizing nozzle that was introduced by the same manufacturer as for nozzle A. To optimize the spatial drug distribution pattern and achieve higher intraabdominal aerosol particle concentration, the conventional axial nozzle is supplemented in nozzle D by three further nozzles, which are arranged lateral at the nozzle head with an angular distance of 120°. Nozzle D is based on the same technology as nozzle A. At the manufacturer-recommended flow rate of 1.5 ml/s, nozzle D shows an operational pressure of 16.0 bar after an initiation time of 92 s and provides four full spray cone jets (67°) composed of droplets with a mass median diameter of 21 μm.

Recently, a first attempt regarding recommendations on the minimum technical requirements on nozzles suitable for PIPAC treatment was published. A minimum requirement for the spray angle of at least 70° was defined [12] by implying that the spray cone angle corresponds to the achievable drug deposition area. In this study we found that not all nozzles fulfil this requirement. The requirement is matched by nozzles A, B and C. Nozzle D achieves a slightly lower spray cone angle (67°) than required, however the four spatially-displaced spray jets achieve a total spray angle of 268°. Nozzle C, unlike all other nozzles examined, produces a hollow spray cone, resulting in a ring-shaped drug deposition area that was smaller than that of a full spray cone jet at the same spray cone angle. Regarding nozzle performance, the drug deposition area seems to be an even better technical parameter than the spray cone angle.

The nozzles C (MCR-4 TOPOL®) and D (QuattroJet) investigated in this study, in contrast to A (Nebuliser) and B (HurriChemTM), and thus in contrast to the primary PIPAC nozzle technology, show significant differences in their operating principle and performance. Nozzle C offers the largest spray cone angle of all examined nozzles, but produces a hollow spray cone. It is not known if such a spray jet improves drug distribution and drug penetration, since no preclinical studies exist comparing a hollow with a full spray cone. Nozzle D provides multiple spray cones that can significantly improve the spatial drug distribution by reduction of high local deposition and thus high local tissue toxicity. Nonetheless, these potential benefits of multi-nozzle systems need to be confirmed by further research.

In contrast to the other nozzles, nozzle C (MCR-4 TOPOL®) exhibits a macroscopically visible formation of iron oxide after long-term exposure to a cytostatic solution containing sodium chloride. Based on spark optical emission spectrometry, it was discovered that nozzle C was made from steel with a significantly higher sulphur content (0.012 wt.-% vs. 0.001 wt.-%), along with contamination of molybdenum (0.183 wt.-%), copper (0.220 wt.-%), and tungsten (0.134 wt.-%). Short-term exposure of the nebulizers to cytostatic solution reveals no immediate macroscopic corrosion. Nevertheless, it is not possible to completely rule out a potential risk to the patient.

Currently, there are only limited preclinical data to suggest that there is an optimal technique for the generation and delivery of PIPAC aerosols that could improve clinical outcome. However, it is clear that, contrary to claims made by one manufacturer [13], larger aerosol droplets injected into the peritoneal cavity at higher velocities with a hollow spray cone do neither improve the spatial distribution pattern [8, 14] nor tissue penetration depth per se. Nebulizers differing from the present standard technology in design, especially in spraying characteristics, cannot automatically be considered equivalent by the clinical user. Therefore, before their broad clinical use, the individual innovation phases should be systematically tested, ideally following the recommendation of the IDEAL-D framework for the introduction of medical devices [15, 16]. While the original nozzle technology has completed phase I - IIb [17] and phase III trials are ongoing [4, 5], only limited phase I clinical user data has been published for nozzle C (MCR-4 Topol^®^) [18] and such data are lacking for nozzle B and D.

In the near future, clinical users should have the assurance that technical testing and reporting adhere to scientific standards and widely accepted global standards, including ISO standards. For PIPAC nebulizers, such standards should ideally be set by a panel of experts. Moreover, nebulizers with significant technical and granulometric differences from the standard technology should undergo first ex- and in-vivo animal testing before clinical use. Manufacturers should be obliged to have the bioequivalence of the cytostatic drugs administered independently certified in comparison to standard nebulizer systems, analogous to drugs and their generics. Relevant outcome measures are aerosol characteristics, spatial drug distribution, depth of penetration, tissue concentration and the peak concentration and the area under the curve describing the extent of peritoneal passage [8, 19 - 28]. The ratio between the individual properties of the generic nebulizer and the reference product would ideally be 1:1 in case of bioequivalence. As this is unlikely to be achieved, the US Food and Drug Administration (FDA), for example, requires the 90% confidence interval for drugs and their generics to be between 0.80 and 1.25 [29]. Similar to such FDA specifications, new PIPAC nozzle technologies could be tested comparatively in the future. Such preclinical testing, ideally using standardized models, could prevent the use of devices compromising clinical outcomes and/or harming healthcare professionals/patients. Finally, it would be helpful for the comparability of clinical results if the nebulizer type used in each case will be also recorded in the PIPAC database (https://isspp.org/professionals/pipac-database/).

## 5 Conclusion

Four clinically-used nozzles to aerosolise chemotherapeutic drugs in the context of Pressurized Intraperitoneal Aerosol Chemotherapy (PIPAC), i.e., the Nebulizer, the HurriChem™, the MCR-4 TOPOL^®^ and the QuattroJet were comparatively examined to determine their performance.

We confirmed that the Nebulizer exhibits a nearly identical technical design, and thus similar performance to the original PIPAC nozzles MIP/CapnoPen. The PIPAC nozzle HurriChem™ is based on a similar technical design as the Nebulizer nozzle but provides a finer aerosol due to a smaller nozzle orifice opening. Both, the MCR-4 TOPOL^®^ and the QuattroJet deviate in the principles of design and operations from the Nebulizer, and thus, the original PIPAC technology. While the MCR-4 TOPOL^®^ provides the coarsest aerosol of the four nozzles examined, the QuattroJet delivers an aerosol similar to that of the HurriChem™. In contrast to the HurriChem™, the QuattroJet comes with the feature of four spray cones (one axial, three lateral) to improve the spatial drug distribution and a higher aerosol particle number concentration.

The availability of new PIPAC nozzles with unique features is encouraging but can also negatively impact optimization and standardization of PIPAC protocols for the treatment of peritoneal carcinomatosis. It is therefore recommended that nozzles for which the technical/granulometric characteristics differ from the current standard technology must be subjected to preclinical proof of equivalence in terms of spatial drug distribution, tissue penetration and concentration before routine clinical use. New nebulizers should be investigated and introduced for clinical use in accordance with the IDEAL-D framework.

## Data Availability

All data produced in the present work are contained in the manuscript

## Acknowledgments

The authors thank Professor Marc Pocard, Hepato-Biliary-Pancreatic Gastrointestinal Surgery and Liver Transplantation, Pitié Salpêtrière Hospital, AP-HP, F-75013 Paris, France, for providing a HurriChem™ (ThermaSolutions, White Bear Lake, MN, USA) nozzle.

## Notes

**Disclosure** Strictly academic study supported by institutional funds. All authors have no conflicts of interest or financial ties to declare.

### Competing Interest Statement

The authors have declared no competing interest.

### Funding Statement

This study did not receive any funding

### Summary of Updates

A linguistic revision of the manuscript took place. Data, tables and images were not modified. Likewise, the technical content - or the data interpretation - has not been changed.

